# Influenza Mortality as an Indicator of the Efficacy of COVID-Related, Non-pharmaceutical Interventions to Reduce the Spread of Respiratory Infections

**DOI:** 10.1101/2023.11.30.23299157

**Authors:** Robert Morris

## Abstract

**Background:** Non-pharmaceutical interventions (NPIs) have been criticized as ineffective in preventing COVID. Because it is a new disease with NPIs introduced almost immediately, we have no way to evaluate the counterfactual of non-intervention easily and directly. We do, however, have historic data on influenza, a respiratory disease with similar patterns of transmission and a well-established CDC surveillance system in the US. These data provide us with an excellent way to indirectly evaluate the efficacy of these interventions.

**Results:** During the three seasons prior to COVID from 2016-17 to 2018-19, the mean total US influenza mortality was 9,917 deaths per season. During the pandemic, total influenza mortality declined by 80 percent relative to historical levels. Pediatric influenza morality decreased by 85 percent. At the state level, the average drop in mortality over the two flu seasons of the pandemic was strongly correlated with the percent of the 2020 presidential votes cast for Joe Biden (r^2^=0.39).

**Conclusions:** These data provide strong evidence that COVID NPIs dramatically reduced the spread of influenza. Given its similar routes of transmission, these results support the assertion that these NPIs also substantially reduced COVID transmission, morbidity, and mortality. The effectiveness of NPIs was highly correlated with the political leanings of each state, suggesting that politics influenced the effectiveness of and/or compliance with NPI strategies.

## Background

From the outset of the COVID pandemic, non-pharmaceutical interventions (NPIs) to protect public health have come under heavy criticism for their impact on everything from the economy^1^ to mental health^2–4^ to education^5,6^. Furthermore, almost every intervention has, at some point, been declared ineffective, including masking,^7,8^ school closures,^9^ and business “lockdowns” ^10,11^. One of the challenges in evaluating the effectiveness of NPIs has been that we have no historical data related to COVID, so our means for evaluating interventions were limited. Specifically, we have no historical data to assess the counterfactual of not intervening. Instead, we have been forced to use ecological studies, with all their well-known limitations, and clinical trials, with their inability to easily capture the impact of community-wide interventions.

One way to look at the impact of COVID interventions on the spread of a respiratory pathogen is to consider their impact on a disease for which we do have considerable historical data. Specifically, we can consider data from influenza surveillance. The CDC, for example, has extensive and detailed historical data on morbidity and mortality related to the flu. Multiple studies have noted the dramatic, global decline in influenza incidence^12–14^ and mortality and its association with NPIs.

Focusing on the flu allows us to compare historical patterns of influenza incidence and mortality with those that prevailed during periods when COVID NPIs were in place. Looking at changes in those rates as a percent of pre-COVID rates allows us to use geographic areas as their own controls, eliminating time invariant confounders. We can then consider what factors might have influenced those changes. Given the association of political party with COVID policies, population adherence to those policies, and COVID mortality,^15^ this study evaluates the association between state political preference, as represented by the percent of votes cast for Joe Biden in 2020, with the reduction in influenza mortality during the COVID pandemic.

## Methods

Weekly counts of influenza deaths in both the entire population and in the pediatric population for the period from 2016 through 2023 were abstracted from the CDC FluView System^16^.

Seasonal influenza mortality from each state over the same period were obtained from the same site.

Average annual influenza mortality rates for both populations were calculated for the pre-COVID period 2016 through 2020 and for the two flu seasons during the pandemic, 2020-21 and 2021-22.

At the state level, average flu season mortality was determined for the pre-COVID period and for the two pandemic flu seasons. The decrease in average flu season mortality for each state during the pandemic as compared to average mortality rates prior to the pandemic were calculated for each state.

The percentage of votes in the 2020 election cast for Joe Biden^17^ were used to assess each state’s political leanings during the pandemic. The linear relationship between Democratic leaning and the reduction in influenza mortality during the pandemic was assessed by simple linear regression.

## Results

US total influenza mortality for 2016-2023 is shown in Figure 1. The figure shows dramatic drops in influenza mortality during the 2020-21 and 2021-22 flu seasons. A more normal flu season resumes in 2022-23. The average total mortality for the three pre-pandemic flu seasons was 9,917 deaths per season. In 2020-21, influenza deaths dropped to 931 deaths per season, a 90.6% decrease. In 2021-2022, there were 3,043 flu deaths in the US, higher than the first year of the pandemic, but 69.3% below pre-COVID levels. Overall, during the two years of the pandemic deaths average annual influenza deaths were 1,987 per season or 80% (95% CI 79%-81%) below pre-COVID levels.

**Figure 1.**
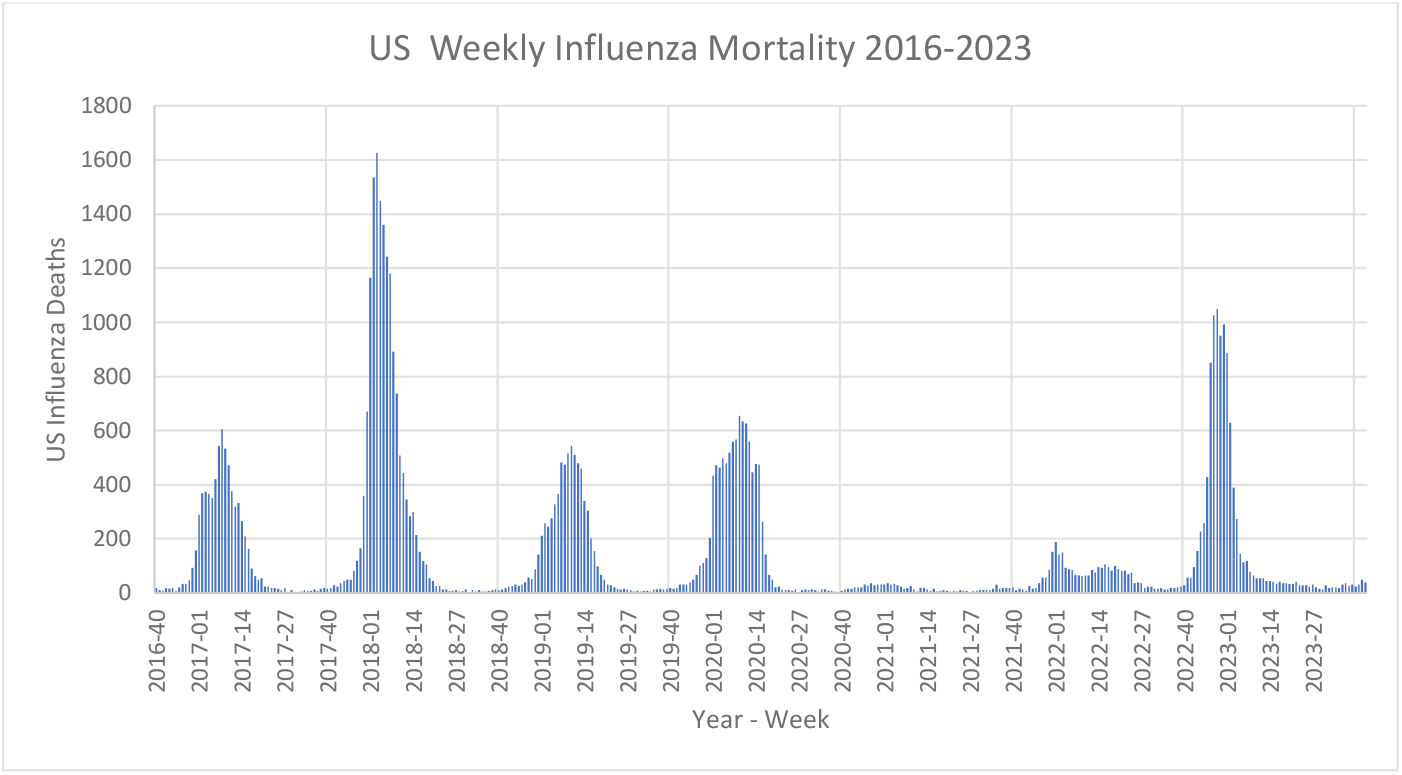
US weekly influenza mortality for the 2016-17 through 2022-23 flu seasons for all ages.

US pediatric influenza mortality is shown in Figure 2. The figure shows dramatic drops in influenza mortality during the 2020-21 and 2021-22 flu seasons. A more normal flu season resumed in 2022-23. The average total influenza mortality for the pre-pandemic flu seasons was 148 deaths per season. In 2020-21, there was only one pediatric influenza death in the entire United States (a 99% decrease from pre-pandemic levels) and rose to 49 deaths per season (a 67% decrease) in 2021-22. Overall, during the two years of the pandemic deaths dropped to 25 per season or 85% (95% CI: 80%-89%) below pre-COVID levels.

**Figure 2.**
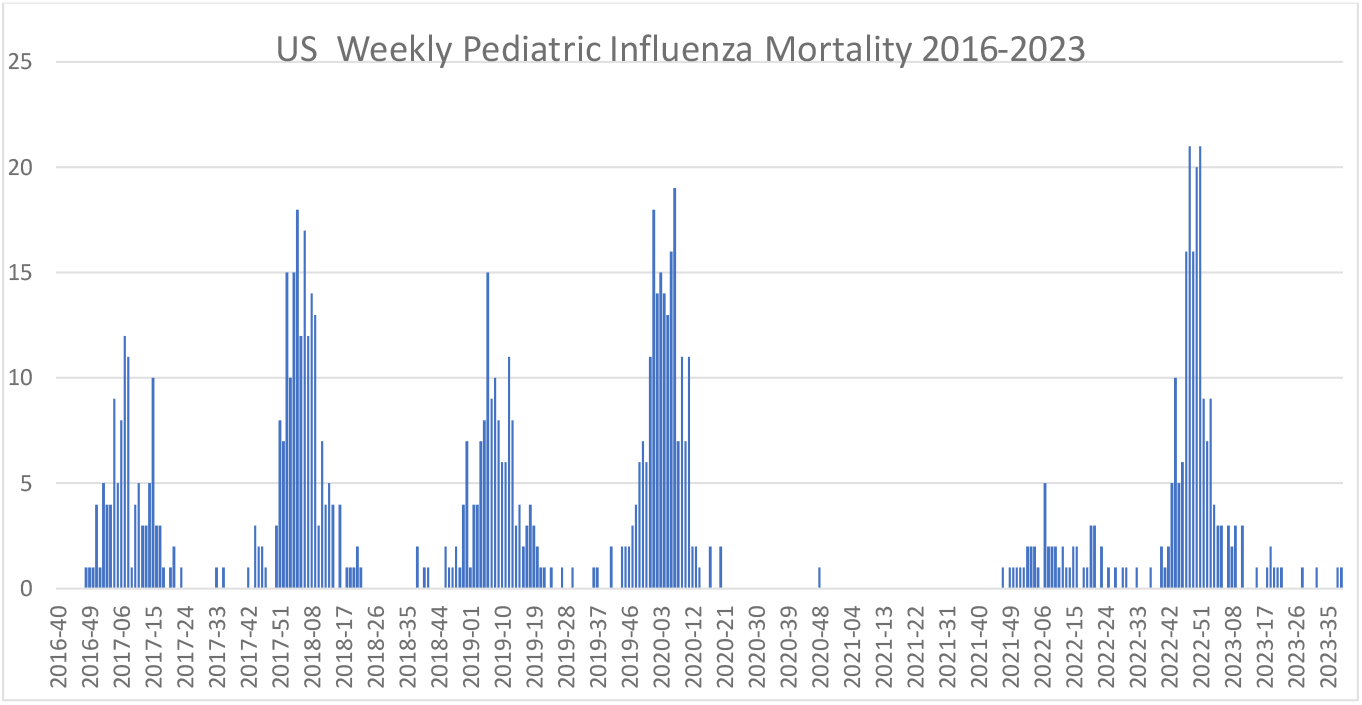
US weekly influenza mortality for the 2016-17 through 2022-23 flu seasons for children.

The total seasonal influenza mortality counts for 2016-2023 are listed in Table 1 for each US state and the District of Columbia along with the percentage of votes cast for Joe Biden in 2020. The table also lists the ratio of seasonal influenza mortality during the pandemic to baseline seasonal influenza mortality. Figure 3 shows the percent decrease in influenza mortality for each as a function of the 2020 Biden vote percentage. The r^2^ of 0.39 is significant at p<0.0001 (based on ANOVA for linear regression).

**Table 1.** Influenza mortality by state and season.

| STATE | 2016-17 | 2017-18 | 2018-19 | 2019-20 | 2020-21 | 2021-22 | Pre Pandemic Average | Pandemic Average | Pandemic Influenza Mortality Decrease | 2020 Biden Vote |
| --- | --- | --- | --- | --- | --- | --- | --- | --- | --- | --- |
| ALABAMA | 60 | 237 | 94 | 154 | 36 | 43 | 130 | 40 | 71.0% | 37% |
| ALASKA | 14 | 27 | 19 | 11 | 1 | 13 | 20 | 7 | 60.6% | 43% |
| ARIZONA | 79 | 329 | 116 | 162 | 9 | 67 | 175 | 38 | 77.8% | 49% |
| ARKANSAS | 54 | 215 | 107 | 108 | 22 | 32 | 125 | 27 | 77.7% | 35% |
| CALIFORNIA | 586 | 1717 | 630 | 897 | 57 | 158 | 978 | 108 | 88.8% | 63% |
| COLORADO | 153 | 249 | 153 | 152 | 6 | 88 | 185 | 47 | 73.4% | 55% |
| CONNECTICUT | 86 | 214 | 90 | 102 | 0 | 29 | 130 | 15 | 88.2% | 59% |
| DELAWARE | 28 | 41 | 23 | 22 | 1 | 6 | 31 | 4 | 87.7% | 59% |
| WASHINGTON DC | 10 | 20 | 8 | 12 | 1 | 0 | 13 | 1 | 96.0% | 92% |
| FLORIDA | 282 | 652 | 341 | 520 | 84 | 218 | 425 | 151 | 66.4% | 48% |
| GEORGIA | 83 | 325 | 114 | 246 | 34 | 91 | 174 | 63 | 67.4% | 49% |
| HAWAII | 38 | 56 | 38 | 29 | 2 | 8 | 44 | 5 | 87.6% | 64% |
| IDAHO | 76 | 114 | 66 | 42 | 6 | 28 | 85 | 17 | 77.2% | 33% |
| ILLINOIS | 216 | 630 | 207 | 249 | 11 | 78 | 351 | 45 | 86.3% | 58% |
| INDIANA | 151 | 444 | 179 | 190 | 16 | 79 | 258 | 48 | 80.3% | 41% |
| IOWA | 143 | 277 | 98 | 116 | 13 | 51 | 173 | 32 | 79.8% | 45% |
| KANSAS | 125 | 241 | 97 | 138 | 11 | 52 | 154 | 32 | 79.0% | 42% |
| KENTUCKY | 127 | 321 | 166 | 163 | 20 | 52 | 205 | 36 | 81.5% | 36% |
| LOUISIANA | 51 | 231 | 64 | 107 | 15 | 40 | 115 | 28 | 75.7% | 40% |
| MAINE | 76 | 88 | 52 | 47 | 4 | 15 | 72 | 10 | 85.6% | 53% |
| MARYLAND | 105 | 171 | 100 | 164 | 9 | 37 | 125 | 23 | 83.0% | 65% |
| MASSACHUSETTS | 198 | 309 | 179 | 203 | 18 | 92 | 229 | 55 | 75.3% | 66% |
| MICHIGAN | 255 | 463 | 239 | 299 | 19 | 111 | 319 | 65 | 79.3% | 51% |
| MINNESOTA | 212 | 328 | 94 | 163 | 11 | 59 | 211 | 35 | 82.4% | 52% |
| MISSISSIPPI | 30 | 166 | 48 | 79 | 34 | 35 | 81 | 35 | 57.3% | 41% |
| MISSOURI | 192 | 522 | 132 | 204 | 12 | 85 | 282 | 49 | 81.5% | 41% |
| MONTANA | 60 | 69 | 36 | 41 | 2 | 22 | 55 | 12 | 76.7% | 41% |
| NEBRASKA | 82 | 114 | 87 | 57 | 5 | 41 | 94 | 23 | 72.9% | 39% |
| NEVADA | 24 | 88 | 43 | 65 | 8 | 26 | 52 | 17 | 69.1% | 50% |
| NEW HAMPSHIRE | 50 | 73 | 47 | 31 | 2 | 21 | 57 | 12 | 77.1% | 53% |
| NEW JERSEY | 115 | 279 | 146 | 178 | 5 | 42 | 180 | 24 | 86.9% | 57% |
| NEW MEXICO | 34 | 80 | 63 | 76 | 5 | 27 | 59 | 16 | 74.7% | 54% |
| NEW YORK | 204 | 478 | 280 | 286 | 31 | 105 | 321 | 68 | 78.2% | 61% |
| NORTH CAROLINA | 310 | 545 | 272 | 305 | 22 | 73 | 376 | 48 | 86.7% | 49% |
| NORTH DAKOTA | 25 | 33 | 20 | 24 | 6 | 20 | 26 | 13 | 49.0% | 32% |
| OHIO | 277 | 662 | 284 | 333 | 18 | 98 | 408 | 58 | 85.1% | 45% |
| OKLAHOMA | 126 | 288 | 135 | 158 | 36 | 91 | 183 | 64 | 64.1% | 32% |
| OREGON | 246 | 259 | 200 | 96 | 6 | 23 | 235 | 15 | 92.8% | 56% |
| PENNSYLVANIA | 334 | 644 | 361 | 344 | 30 | 159 | 446 | 95 | 77.5% | 50% |
| RHODE ISLAND | 49 | 62 | 46 | 32 | 2 | 9 | 52 | 6 | 88.4% | 59% |
| SOUTH CAROLINA | 119 | 313 | 138 | 166 | 18 | 48 | 190 | 33 | 82.1% | 43% |
| SOUTH DAKOTA | 44 | 63 | 39 | 29 | 4 | 27 | 49 | 16 | 64.6% | 36% |
| TENNESSEE | 142 | 401 | 165 | 262 | 60 | 106 | 236 | 83 | 65.8% | 37% |
| TEXAS | 357 | 1165 | 448 | 640 | 148 | 260 | 657 | 204 | 68.7% | 46% |
| UTAH | 51 | 98 | 63 | 58 | 6 | 28 | 71 | 17 | 74.8% | 38% |
| VERMONT | 22 | 48 | 35 | 24 | 1 | 5 | 35 | 3 | 90.7% | 66% |
| VIRGINIA | 144 | 292 | 180 | 167 | 23 | 59 | 205 | 41 | 79.1% | 54% |
| WASHINGTON | 380 | 409 | 292 | 196 | 6 | 35 | 360 | 21 | 93.6% | 58% |
| WEST VIRGINIA | 46 | 157 | 66 | 73 | 1 | 45 | 90 | 23 | 73.1% | 30% |
| WISCONSIN | 186 | 382 | 126 | 179 | 7 | 55 | 231 | 31 | 85.8% | 49% |
| WYOMING | 14 | 25 | 22 | 11 | 1 | 8 | 20 | 5 | 75.0% | 27% |
| TOTAL | 6871 | 15414 | 7048 | 8410 | 905 | 3000 | 9778 | 1953 | 80.0% |  |

**Figure 3.**
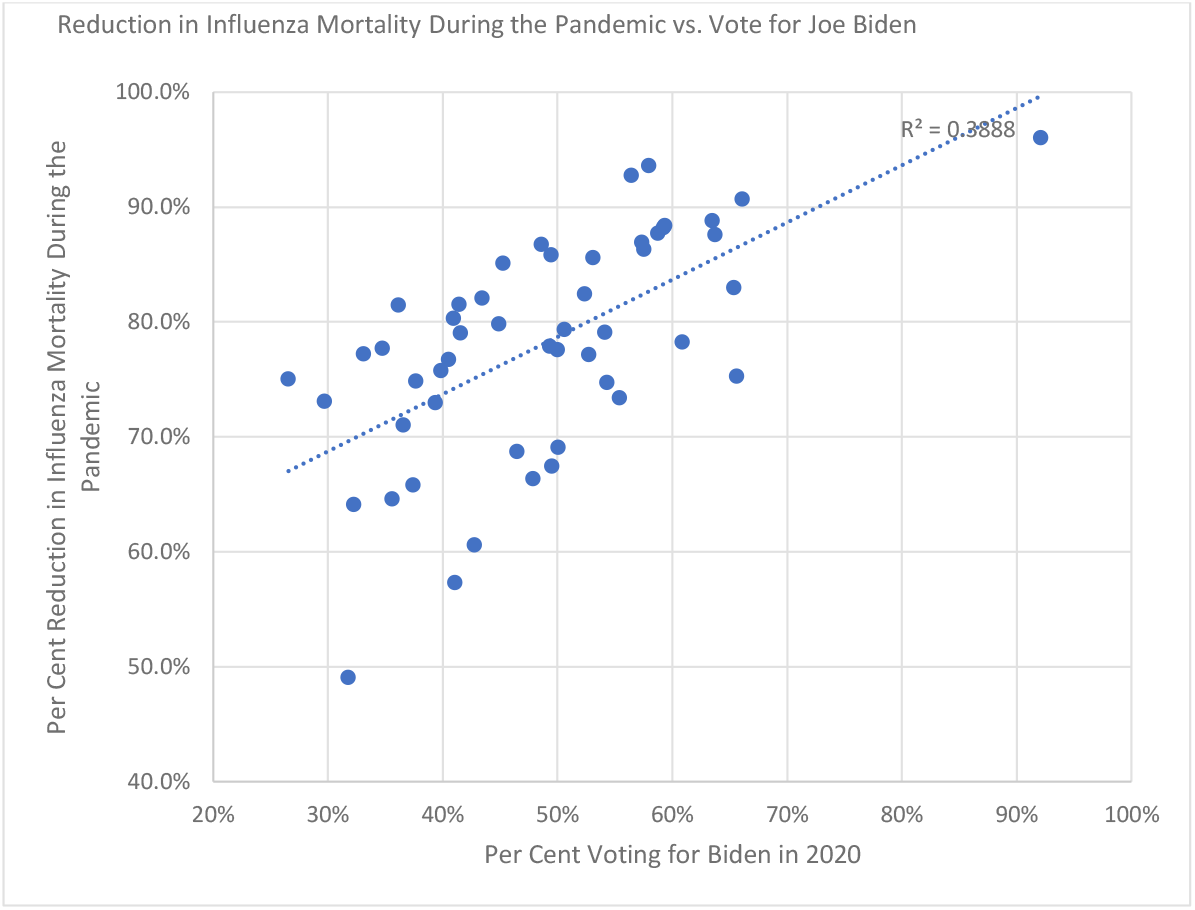
Average annual reduction in influenza during the 2020-21 and 2021-2022 flu seasons for US States as a function of the percent vote for Joe Biden in the 2020 Presidential election.

## Conclusions

The data demonstrate dramatic reductions in US influenza mortality during the COVID pandemic. Since we can reject the null hypothesis that these are simply random fluctuations reductions in flu severity, we must conclude that these changes are the result of either unexplained decreases in ascertainment or the introduction of public health measures that reduced transmission. Influenza surveillance and diagnostic tools are extremely well-established by the CDC, so decreased ascertainment, particularly for mortality, seems unlikely. The Seattle Flu Study demonstrates the same pandemic-related decrease in influenza as well as other respiratory viruses, including respiratory syncytial virus, with an active surveillance program.^18,19^

Excluding decreased ascertainment as an explanation means something that occurred during the COVID pandemic reduced the spread of respiratory viruses. The most likely explanation is that NPIs imposed to reduce the spread of COVID were extremely effective in reducing the transmission of respiratory viruses, particularly influenza. This effect was most dramatic during the 2020-21 flu season when the most stringent NPIs were in place in the United States. These measures were relaxed in 2021-2022, and influenza mortality rose, but was still far below pre-COVID levels. Overall, these data suggest that almost 16,000 influenza deaths were prevented in the United States alone when COVID NPIs were in place. WHO FluNet data show similar reductions worldwide in influenza during the pandemic.^20^

This effect was more dramatic in children than adults, with the near complete elimination of pediatric influenza mortality in 2020-21, from an average of 148 deaths pre-COVID to a single death during the first full flu season during COVID. The greater mortality reduction in children (>99% vs 90%) suggests interventions in children were even more effective than interventions in adults. The major difference in NPIs for children as opposed to adults is that school closures tended to be more universal than business closures, particularly during the 2020-21 school year.

It seems likely that those NPIs produced similar percentage reductions in COVID mortality given similar patterns of transmission. Even a 50% reduction in COVID mortality would correspond to 1.1 million lives saved by NPIs.

Implementation of NPIs and the population adherence to NPIs differed widely from state to state, driven, in large part, by politics. The strong correlation of the drop in influenza mortality with the Democratic Presidential vote in 2020 suggests that political leanings of each state influenced the efficacy of NPIs. Other studies have shown associations between political leanings and both the utilization and effectiveness of NPIs for COVID.^21,22^ These results provide compelling evidence which eliminates time invariant confounders.

In sum, COVID NPIs appear to have been highly effective in reducing the spread of respiratory infections, with the magnitude of that benefit associated with each state’s political leanings. We cannot determine with confidence which NPIs are responsible with this data set, but it is essential that we do so to be better prepared to face future pandemics. Any assertion that NPIs were ineffective in reducing the spread of COVID must explain how the spread of COVID is dramatically different from the spread of influenza.

## Data Availability

All data are publicly available through existing websites.

